# Management and Outcomes of Critically-Ill Patients with COVID-19 Pneumonia at a Safety-net Hospital in San Francisco, a Region with Early Public Health Interventions: A Case Series

**DOI:** 10.1101/2020.05.27.20114090

**Authors:** Sky Vanderburg, Narges Alipanah, Rebecca Crowder, Christina Yoon, Richard Wang, Neeta Thakur, Kristin Slown, Priya B. Shete, Martin Rofael, John Z. Metcalfe, Cindy Merrifield, Carina Marquez, Katherine Malcolm, Michael Lipnick, Vivek Jain, Antonio Gomez, Gregory Burns, Lillian B. Brown, Christopher Berger, Vincent Auyeung, Adithya Cattamanchi, Carolyn M. Hendrickson

## Abstract

**Background:** Following early implementation of public health measures, San Francisco has experienced a slow rise and a low peak level of coronavirus disease 2019 (COVID-19) cases and deaths.

**Methods and Findings:** We included all patients with COVID-19 pneumonia admitted to the intensive care unit (ICU) at the safety net hospital for San Francisco through April 8, 2020. Each patient had ≥15 days of follow-up. Among 26 patients, the median age was 54 years (interquartile range, 43 to 62), 65% were men, and 77% were Latinx. Mechanical ventilation was initiated for 11 (42%) patients within 24 hours of ICU admission and 20 patients (77%) overall. The median duration of mechanical ventilation was 13.5 days (interquartile range, 5 to 20). Patients were managed with lung protective ventilation (tidal volume ≤8 ml/kg of ideal body weight and plateau pressure ≤30 cmH_2_O on 98% and 78% of ventilator days, respectively). Prone positioning was used for 13 of 20 (65%) ventilated patients for a median of 5 days (interquartile range, 2 to 10). Seventeen (65%) patients were discharged home, 1 (4%) was discharged to nursing home, 3 (12%) were discharged from the ICU, and 2 (8%) remain intubated in the ICU at the time of this report. Three (12%) patients have died.

**Conclusions:** Good outcomes were achieved in critically ill patients with COVID-19 by using standard therapies for acute respiratory distress syndrome (ARDS) such as lung protective ventilation and prone positioning. Ensuring hospitals can deliver sustained high-quality and evidence-based critical care to patients with ARDS should remain a priority.

## Introduction

Coronavirus infectious disease 2019 (COVID-19) caused by the severe acute respiratory syndrome coronavirus-2 (SARS-CoV-2) is now the leading cause of death in the United States.^1^ The number of reported cases and deaths varies markedly in part due to regional variation in public health responses to the pandemic.^2-5^

Initial reports of outcomes among adults with COVID-19 requiring intensive care unit (ICU) admission showed high overall mortality (62% in China, 26% in Italy, 50% in the United Kingdom, and 50-67% in the US) and mortality exceeding 80% among patients requiring invasive mechanical ventilation.^6-9^ Mortality has been variable in subsequent reports but exceeds 50% in nearly half of published studies.^4,7,10-12^ Many of these reports are from settings that experienced surges of patients that overwhelmed hospital capacity and presented a challenge to delivering standard ICU care.

San Francisco was among the first cities in the country to implement strong physical distancing measures. The Mayor declared a state of emergency on February 25 and a regional shelter-in-place order was issued on March 16.^13,14^ The ordinances occurred 28 days and 8 days, respectively, before the first death from COVID-19 was reported in San Francisco on March 24. These actions enabled healthcare systems to mobilize resources for COVID-19 admissions and likely flattened the epidemic curve at a low peak level (1,312 cases of COVID-19 and 21 deaths in San Francisco as of April 23).^15^

Here, we report the first detailed ICU case series from a healthcare system in the United States with sufficient lead time and capacity to increase staffing of ICU care teams, establish dedicated COVID-19 ICUs, and procure supplies and equipment. The objective is to describe the demographics, clinical characteristics, ICU management and outcomes of critically ill patients with COVID-19 pneumonia at Zuckerberg San Francisco General Hospital and Trauma Center (ZSFG).

## Methods

### STUDY SETTING

ZSFG is a 284-bed hospital (58 ICU beds) operated by the San Francisco Department of Public Health and staffed by physicians from the University of California San Francisco. It is the primary provider of safety-net health care and 80% of its patient population receives publicly funded health insurance or is uninsured. San Francisco is the second most densely populated city in the United States and the hospital’s catchment includes a population of 881,549 within 46.7 square miles.

To date, COVID-19 ICU admissions have been clustered in two 10-bed ICUs. In addition to regular staffing of medical and surgical ICUs, COVID-19 ICUs were staffed by dedicated physician teams (one intensivist, one critical care fellow, and two internal medicine or anesthesia residents per 10 patients), 1:1 nursing with an extra nurse supporting care for every two mechanically ventilated patients, one respiratory therapist (RT) for up to 4 mechanically ventilated patients with an extra RT available on each unit for additional support, and a dedicated ICU pharmacist. Five ICU staff members including at least one RT and two COVID-19 ICU nurses supported prone positioning. A dedicated COVID-19 anesthesia team (one attending physician and one advanced practice provider or resident physician) was available 24 hours a day for intubation bundled with placement of vascular access. During the study period, the hospital did not experience shortages in critical equipment or most drugs, and at peak ICU census, 43% of available ventilators were in use. Adequate personal protective equipment and infection control measures were in place (only one clinical staff member has tested positive for SARS-CoV-2 to date).

### STUDY POPULATION AND DATA COLLECTION

All adults (18 years of age or older) admitted to the ICU with respiratory failure between March 24 and April 8 and confirmed to have COVID-19 by an internally-validated reverse transcription polymerase chain reaction assay were included. The UCSF Committee on Human Research approved the study and waived the requirement for informed consent (researchers analyzed only de-identified, anonymized data).

Data from the electronic medical record were extracted using a form in Research Electronic Data Capture software (REDCap).^16,17^ We obtained demographic data, information on clinical symptoms or signs at presentation, results of laboratory and radiologic studies, co-existing conditions, daily ICU management data, and patient outcomes.

### STUDY DEFINITIONS

Daily ventilator parameters, use and duration of mechanical ventilation, prone positioning and neuromuscular blockade were collected through maximum 28 days of follow up. Patient outcome data including hospitalization status were updated through May 10. Acute respiratory distress syndrome (ARDS) was defined using the Berlin criteria.^18^

### STATISTICAL ANALYSIS

Descriptive statistics were used to summarize the data; results are reported as medians and interquartile ranges or means and standard deviations, as appropriate. Categorical variables were summarized as counts and percentages. Median and individual values for selected ventilator parameters were plotted for each day of mechanical ventilation through 14 days of follow up. Analysis was performed with Stata 15.1 software (StataCorp).^19^

## Results

### PATIENT DEMOGRAPHIC AND CLINICAL CHARACTERISTICS

The median age of the study population was 54 years (interquartile range, 43 to 62), 35% were women, 77% identified as Latinx, and 12% had marginal housing (**Table 1**). The median BMI was 31.4 (interquartile range, 27.8 to 35.3), 20 (77%) had a documented comorbidity prior to hospitalization, and 12 (46%) had at least two comorbidities. The most prevalent comorbidities were diabetes mellitus (n = 15, 58%) with a median hemoglobin A1c of 9.4 (interquartile range, 8.0 to 11.8) and hypertension (n = 14, 54%).

**Table 1.**
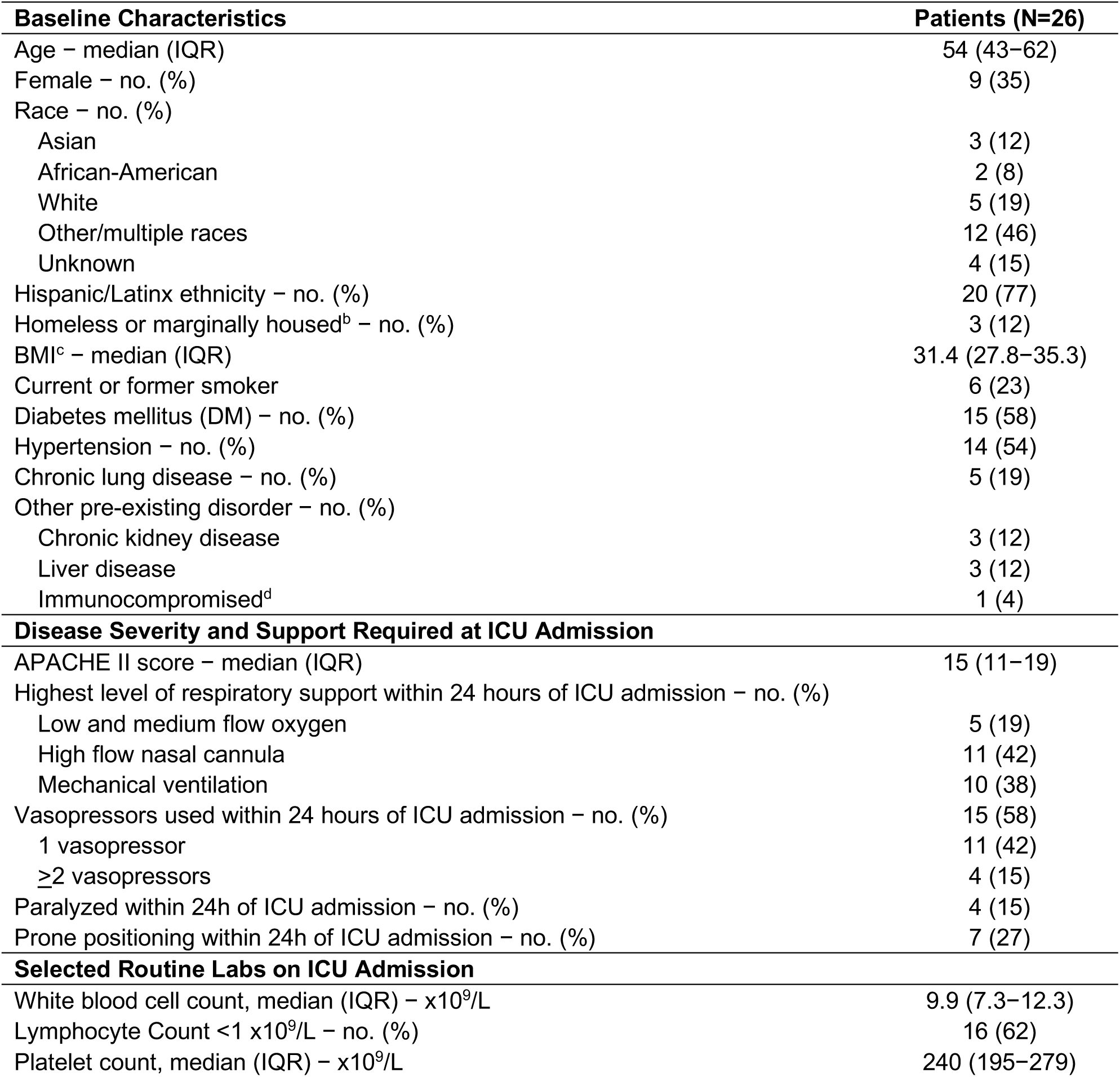
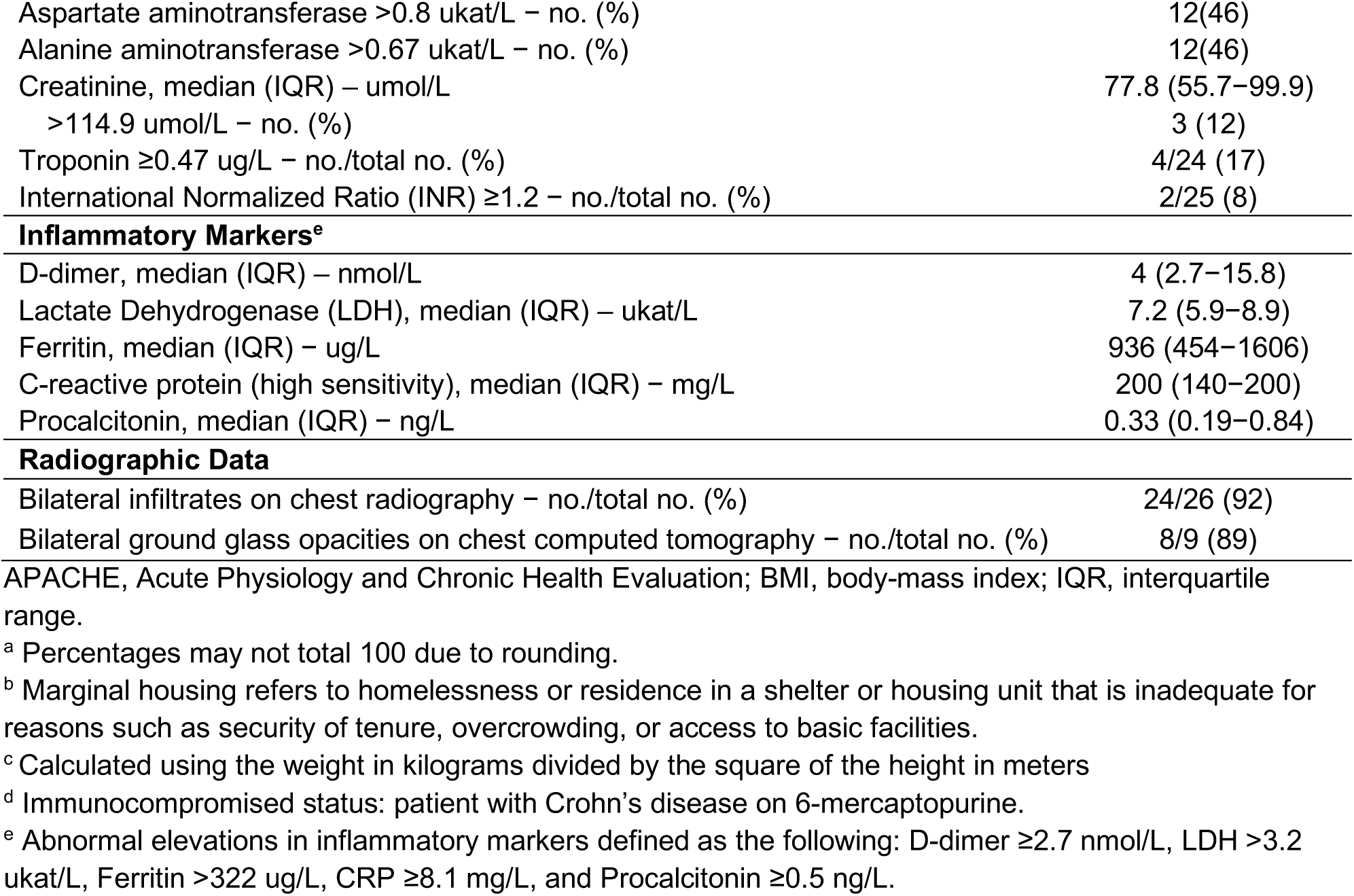
Characteristics of Patients at Baseline and at ICU Admissions.^a^.

| Baseline Characteristics | Patients (N=26) |
| --- | --- |
| Age – median (IQR) | 54 (43–62) |
| Female – no. (%) | 9 (35) |
| Race – no. (%) |  |
| Asian | 3 (12) |
| African-American | 2 (8) |
| White | 5 (19) |
| Other/multiple races | 12 (46) |
| Unknown | 4 (15) |
| Hispanic/Latinx ethnicity – no. (%) | 20 (77) |
| Homeless or marginally housed<sup>b</sup> – no. (%) | 3 (12) |
| BMI<sup>c</sup> – median (IQR) | 31.4 (27.8–35.3) |
| Current or former smoker | 6 (23) |
| Diabetes mellitus (DM) – no. (%) | 15 (58) |
| Hypertension – no. (%) | 14 (54) |
| Chronic lung disease – no. (%) | 5 (19) |
| Other pre-existing disorder – no. (%) |  |
| Chronic kidney disease | 3 (12) |
| Liver disease | 3 (12) |
| Immunocompromised<sup>d</sup> | 1 (4) |
| **Disease Severity and Support Required at ICU Admission** |  |
| APACHE II score – median (IQR) | 15 (11–19) |
| Highest level of respiratory support within 24 hours of ICU admission – no. (%) |  |
| Low and medium flow oxygen | 5 (19) |
| High flow nasal cannula | 11 (42) |
| Mechanical ventilation | 10 (38) |
| Vasopressors used within 24 hours of ICU admission – no. (%) | 15 (58) |
| 1 vasopressor | 11 (42) |
| ≥2 vasopressors | 4 (15) |
| Paralyzed within 24h of ICU admission – no. (%) | 4 (15) |
| Prone positioning within 24h of ICU admission – no. (%) | 7 (27) |
| **Selected Routine Labs on ICU Admission** |  |
| White blood cell count, median (IQR) – x10<sup>9</sup>/L | 9.9 (7.3–12.3) |
| Lymphocyte Count <1 x10<sup>9</sup>/L – no. (%) | 16 (62) |
| Platelet count, median (IQR) – x10<sup>9</sup>/L | 240 (195–279) |
| Aspartate aminotransferase >0.8 ukat/L – no. (%) | 12(46) |
| Alanine aminotransferase >0.67 ukat/L – no. (%) | 12(46) |
| Creatinine, median (IQR) – umol/L | 77.8 (55.7–99.9) |
| >114.9 umol/L – no. (%) | 3 (12) |
| Troponin ≥0.47 ug/L – no./total no. (%) | 4/24 (17) |
| International Normalized Ratio (INR) ≥1.2 – no./total no. (%) | 2/25 (8) |
| Inflammatory Markers<sup>e</sup> |  |
| D-dimer, median (IQR) – nmol/L | 4 (2.7–15.8) |
| Lactate Dehydrogenase (LDH), median (IQR) – ukat/L | 7.2 (5.9–8.9) |
| Ferritin, median (IQR) – ug/L | 936 (454–1606) |
| C-reactive protein (high sensitivity), median (IQR) – mg/L | 200 (140–200) |
| Procalcitonin, median (IQR) – ng/L | 0.33 (0.19–0.84) |
| Radiographic Data |  |
| Bilateral infiltrates on chest radiography – no./total no. (%) | 24/26 (92) |
| Bilateral ground glass opacities on chest computed tomography – no./total no. (%) | 8/9 (89) |

APACHE, Acute Physiology and Chronic Health Evaluation; BMI, body-mass index; IQR, interquartile range.
<sup>a</sup> Percentages may not total 100 due to rounding.
<sup>b</sup> Marginal housing refers to homelessness or residence in a shelter or housing unit that is inadequate for reasons such as security of tenure, overcrowding, or access to basic facilities.
<sup>c</sup> Calculated using the weight in kilograms divided by the square of the height in meters
<sup>d</sup> Immunocompromised status: patient with Crohn's disease on 6-mercaptopurine.
<sup>e</sup> Abnormal elevations in inflammatory markers defined as the following: D-dimer ≥2.7 nmol/L, LDH >3.2 ukat/L, Ferritin >322 ug/L, CRP ≥8.1 mg/L, and Procalcitonin ≥0.5 ng/L.

Patients experienced symptoms for a median of 8 days (interquartile range, 5 to 13) prior to ICU admission (**Figure 1**). Eleven (42%) patients were directly admitted to the ICU of whom 4 required immediate mechanical ventilation, and 7 were placed on high flow nasal cannula. Of 15 patients transferred to the ICU from an acute care bed, 11 (73%) were intubated within 24 hours of transfer. The median APACHE II score was 15 (interquartile range, 11 to 19) at the time of ICU admission (**Table 1**).^20^ Twenty (77%) patients met the Berlin criteria for ARDS with 19 categorized as having moderate to severe disease based on the ratio of the partial pressure of arterial oxygen to the fraction of inspired oxygen [PaO_2_:FiO_2_] <200.

**Figure 1.**
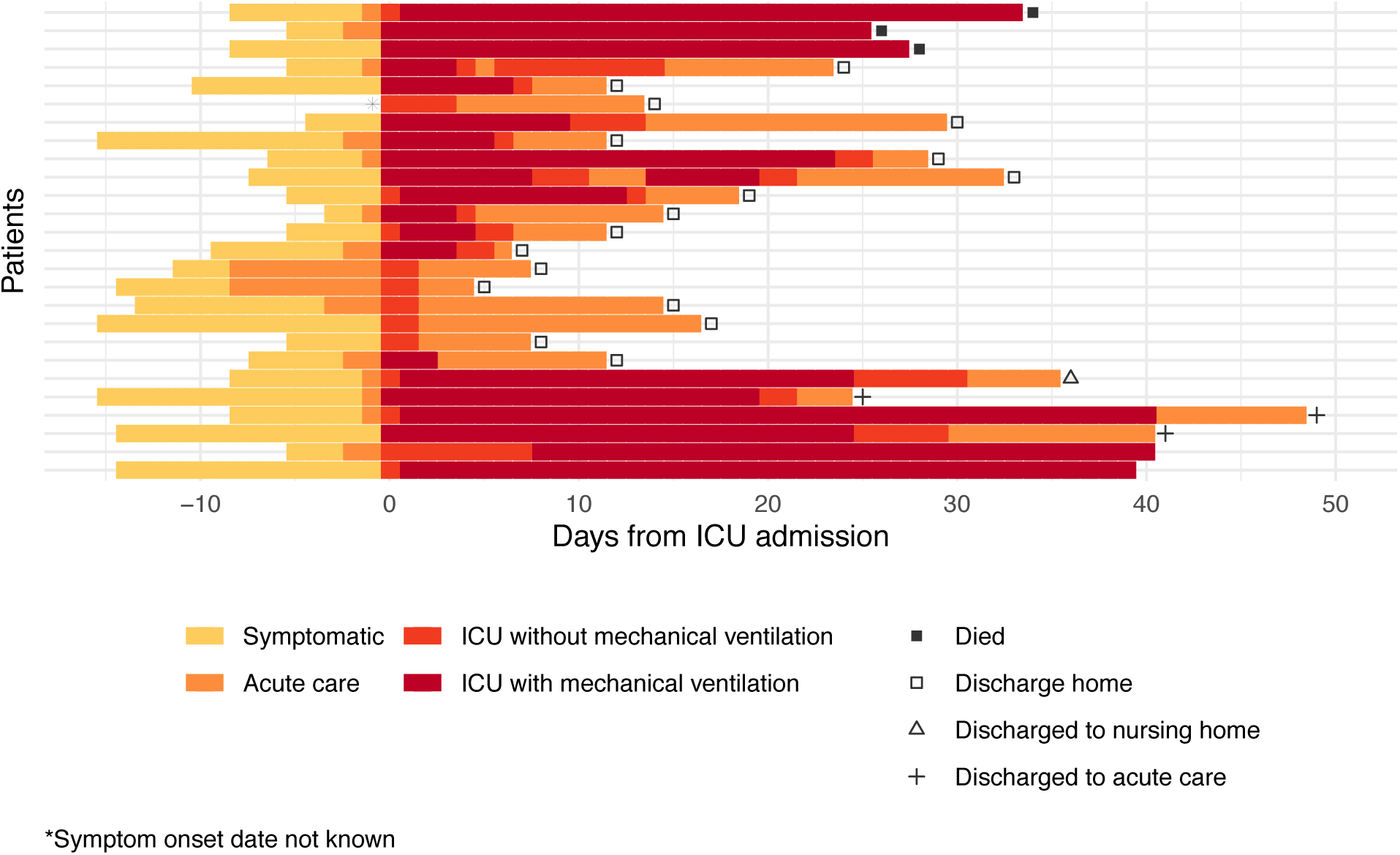
Patient Clinical Course. Clinical course is depicted from symptom onset as reported by the patient on admission through termination of data collection on May 10, 2020. Patients had symptoms for a median duration of 8 days (IQR 5-13) before intensive care unit (ICU) admission. Fifteen (58%) patients were hospitalized for a median duration of 2 days (IQR 1-2) before requiring ICU level care and 11 (42%) were admitted directly to the ICU. Twenty (77%) patients required mechanical ventilation for a median duration of 13.5 days (IQR 5-20). As of May 10, 2020, 17 (65%) patients had been discharged home, 1 (4%) had been discharged to a skilled nursing facility, 3 (12%) had been transferred from ICU to acute care, and 2 (8%) remained on mechanical ventilation. Three patients (12%) had died. Unless otherwise shown, the ICU admission is ongoing.

### LABORATORY, RADIOLOGIC, AND MICROBIOLOGIC FINDINGS

The white blood cell count (WBC) was normal for most patients (median 9.9 ×10^9^/L; interquartile range, 7.3 to 12.3), though leukocytosis (WBC >12 ×10^9^/L) and lymphopenia (<1 ×10^9^/L) were present in 31% and 62% of patients, respectively (**Table 1**). Transaminitis was present in 46% of patients. Most patients had elevated blood inflammatory markers, including D-dimer (74%), lactate dehydrogenase (95%), ferritin (92%), and C-reactive protein (100%).

Chest radiographs obtained before ICU admission demonstrated bilateral infiltrates in 24 (92%), unilateral infiltrates in 1 (4%), and no infiltrates in 1 (4%) (**Table 1**). Chest computed tomography revealed bilateral ground glass opacities in 8 (89%) of 9 studies. At the time of ICU admission, there were no viral co-infections among 23 patients tested and no culture-confirmed bacterial co-infections.

### CLINICAL COURSE AND TREATMENT

Six (23%) patients received supplemental oxygen via high flow nasal cannula and 20 (77%) required mechanical ventilation during their ICU course (**Figure 1**). Adherence to lung protective ventilation strategies for patients with ARDS was excellent. The median tidal volume delivered was 6.0 ml/kg of ideal body weight (IBW) (interquartile range, 5.6 to 6.8) and remained ≤6 ml/kg IBW on 72% of ventilator days and ≤8 ml/kg IBW on 98% of ventilator days. The median plateau pressure on admission was 25 cmH_2_O (interquartile range, 22 to 28) and was ≤30 cmH_2_O on 78% of ventilator days. Among those receiving mechanical ventilation, daily ventilator parameters showed marked patient-to-patient variation (**Figure 2**). The median severity of ARDS was moderate at the onset of mechanical ventilation (PaO_2_:FiO_2_ 159, interquartile range, 104 to 217). The median lung compliance was low at the onset of mechanical ventilation (31 mL/cmH_2_O, interquartile range, 23 to 39, normal is 50-100 mL/cmH_2_O) and stayed low over 14 days of follow-up. The median plateau pressure was ≤25 cmH_2_O on each of the first five days of mechanical ventilation and increased to 25 to 30 cmH_2_O thereafter. Five patients (25%) had driving pressures ≥14 cmH_2_O on the first day of mechanical ventilation and 65% of intubated patients had a driving pressure ≥14 cmH_2_O on at least one day during follow-up.

**Figure 2.**
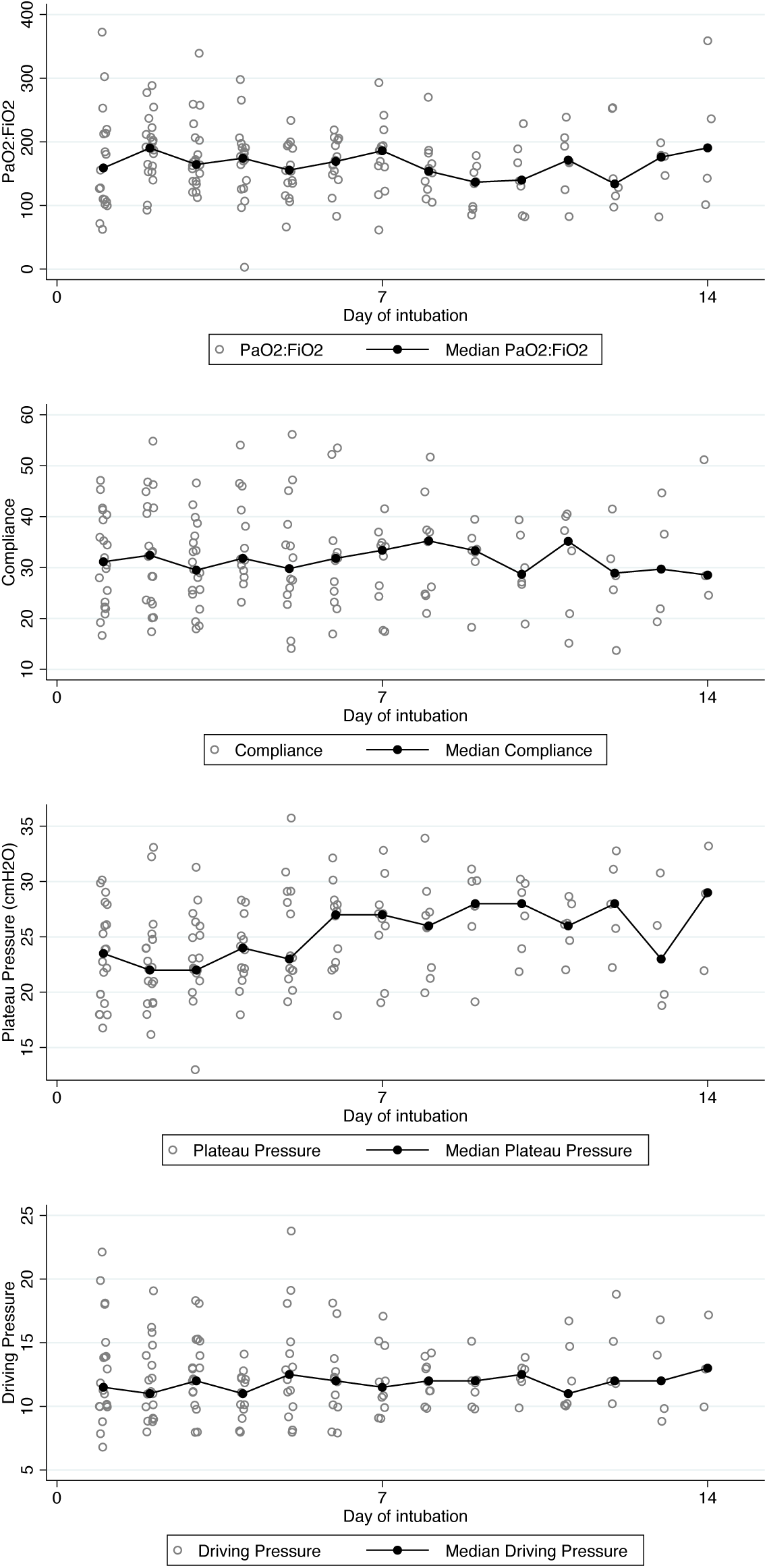
Respiratory Metrics Among Mechanically Ventilated Patients. Each white dot represents the daily value recorded closest to 8 AM for each patient on mechanical ventilation. The black dot represents the median value for all measurements on a given day of ventilation. PaO_2_ denotes partial pressure of arterial oxygen and FiO_2_ denotes the fraction of inspired oxygen. Compliance is calculated using tidal volume divided by the difference between the plateau and the positive end expiratory pressures. The driving pressure is the difference between the plateau pressure and the positive end expiratory pressure.

In addition to lung protective ventilation, ARDS was primarily managed using standard therapies for hypoxemic respiratory failure (**Figure 3**). In particular, 13 of 20 (65%) patients mechanically ventilated were placed in the prone position for 16-20 hours a day for a median duration of 5 days (interquartile range, 2 to 10) and a maximum of 17 days (**Table 2**). Prone positioning was initiated early in moderate to severe ARDS.^21^ At the time of initiation, the median PaO_2_ was 71 mmHg (interquartile range, 63 to 84), median FiO_2_ was 0.7 (interquartile range, 0.5 to 0.7) and median PEEP was 12 cmH_2_O (interquartile range, 12 to 14). Twelve (46%) patients received neuromuscular blockade for a median duration of 9 days (interquartile range, 6 to 12). Four (15%) patients received inhaled epoprostenol (median 11 days, interquartile range 3 to 12). Mechanically ventilated patients required a high level of sedation (**Table 2**).

**Figure 3.**
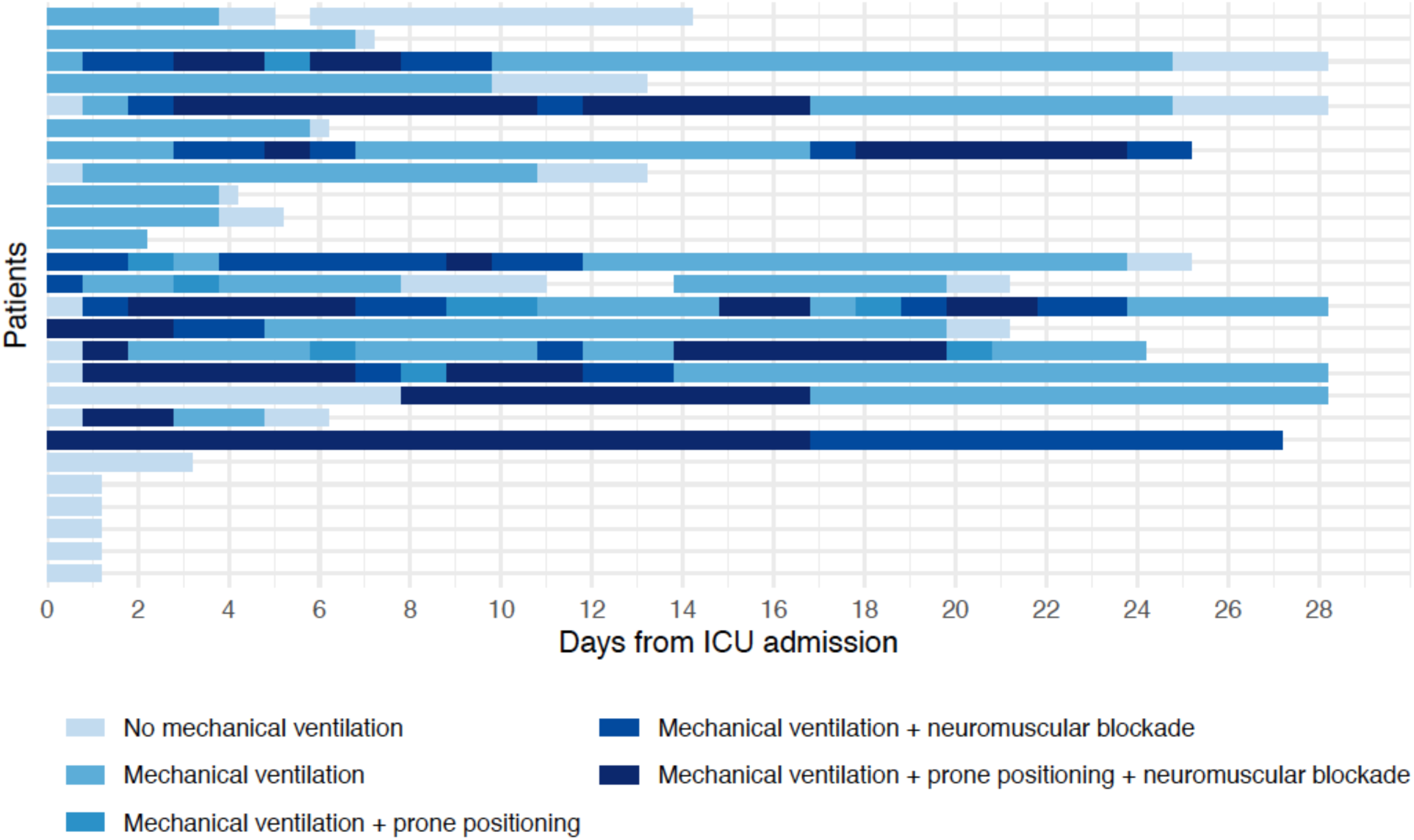
Ventilation Timeline, 24 March-4 May 2020. Mechanically ventilated patients were managed using a lung protective ventilation strategy based on the ARDSNet protocol. Daily ventilation status with or without adjunctive treatments for hypoxemic respiratory failure including prone positioning and neuromuscular blockage are shown through May 4. Breaks correspond to days that a patient was not in the ICU.

**Table 2.** ICU Management and Outcomes.^a^.

| **Treatment of Hypoxemic Respiratory Failure** | **Patients (N=26)** |
| --- | --- |
| Mechanical ventilation – no. (%) | 20 (77) |
| Median duration of mechanical ventilation (IQR) – days | 13.5 (5–20) |
| Prone positioning – no./total no. (%) | 13/20 (65) |
| Median duration of prone positioning (IQR) – days | 5 (2–10) |
| Neuromuscular blockade Infusion - no./total no. (%) | 12/20 (60) |
| Median duration of neuromuscular blockade infusion (IQR) - days | 8.5 (6.5–12) |
| Inhaled epoprostenol – no./total no. (%) | 4/20 (20) |
| Median duration of inhaled epoprostenol (IQR) – days | 11 (3–12) |
| Referred for ECMO consideration – no./total no. (%) | 4/20 (20) |
| Transferred for ECMO – no./total no. (%) | 2/20 (10) |
| **Drug Therapies<sup>b</sup>** |  |
| Remdesivir placebo-controlled trial – no. (%) | 12 (46) |
| Mesenchymal stem cells placebo-controlled trial – no. (%) | 5 (19) |
| Hydroxychloroquine – no. (%) | 13 (50) |
| Median duration of hydroxychloroquine (IQR) – days | 2 (1–5) |
| Azithromycin – no. (%) | 20 (77) |
| Median duration of azithromycin (IQR) – days | 4 (2.5–5) |
| Systemic steroids – no. (%) |  |
| Given for ARDS | 0 (0) |
| Given for other indications | 3 (12) |
| **Sedatives and Analgesic Infusions** |  |
| Propofol - no. (%) | 20 (77) |
| Median of maximum dose (IQR) mcg/kg/min | 80 (80–80) |
| Median duration of infusion (IQR) – days | 8 (3–12) |
| Midazolam – no. (%) | 11 (42) |
| Median of maximum dose (IQR) mg/hr | 5 (4–8) |
| Median duration of infusion (IQR) – days | 9 (3–11) |
| Dexmedetomidine – no. (%) | 14 (54) |
| Median of maximum dose (IQR) mcg/kg/hr | 1.1 (0.7–1.5) |
| Median duration of infusion (IQR) – days | 2 (1–4) |
| Fentanyl – no. (%) | 20 (77) |
| Median of maximum dose (IQR) mcg/hr | 250 (150–350) |
| Median duration of infusion (IQR) – days | 8 (4–14) |
| Hydromorphone – no. (%) | 3 (12) |
| Range of Maximum Infusion – mg/hr | 4–6 |
| Range of duration of infusion – days | 1–19 |
| **Other Therapies** |  |
| Mediation duration of vasopressor use (IQR) – days | 6 (4–13.5) |
| Continuous renal replacement therapy – no. (%) | 5 (19) |
| **Outcomes** |  |
| Hospitalization status – no. (%) |  |
| Discharged to home | 17 (65) |
| Discharged to skilled nursing facility | 1 (4) |
| Discharged to acute care | 3 (12) |
| Still in ICU – mechanically ventilated | 2 (8) |
| Died | 3 (12) |
| Median length of stay (IQR) – days |  |
| Hospital | 15 (12–25) |
| ICU | 9 (3–24) |
| Other adverse outcomes – no. (%) |  |
| Re-intubated within 48 hours – no./total no. (%) | 1/20 (5) |
| Re-admitted to ICU after discharge | 2 (8) |
| Treated for ventilator-associated pneumonia – no./total no. (%) | 6/20 (30) |
| Catheter-associated urinary tract infections | 1 (4) |
| Confirmed central line infection | 0 (0) |

IQR, interquartile range; ARDS, acute respiratory distress syndrome; ECMO, extracorporeal membrane oxygenation.
<sup>a</sup> Percentages may not total 100 due to rounding.
<sup>b</sup> Two mutually exclusive clinical trials were enrolling patients during the study period. Twelve patients (46%) were enrolled in a trial testing Remdesivir vs. placebo (NCT04257656) for Covid-19 disease, and 5 (19%) were enrolled in a trial testing human Mesenchymal Stromal Stem Cells vs. placebo (NCT03818854) for severe ARDS.

Twenty (77%) patients required vasopressor support for a median duration of 6 days (interquartile range, 4 to 13.5), and 5 (19%) patients needed continuous renal replacement therapy (CRRT). No patients received systemic steroids or other immunosuppressive medications for treatment of COVID-19. Three patients received systemic steroids for other indications (one each for refractory septic shock, stridor, and bronchospasm with underlying obstructive lung disease). Nine patients received therapeutic anticoagulation: 2 for confirmed venous thromboembolic disease, 1 to prevent clotting of the CRRT circuit, and 6 for empiric treatment of pulmonary embolism. Anti-coagulation was discontinued in 5 of 6 patients treated empirically after negative imaging studies, and the sixth patient was not stable enough to obtain imaging. Other treatments are summarized in **Table 2**.

### OUTCOMES

Patients were followed for at least 21 hospital days. As of May 10, among the 26 patients, 17 (65%) were discharged home, 1 (4%) was discharged to a skilled nursing facility, 3 (12%) were discharged to an acute care bed, and 2 (8%) were still in the ICU requiring mechanical ventilation (**Table 2**). Three (12%) patients have died.

The median length of hospital admission was 15 days (interquartile range, 12 to 25), and the median length of ICU admission was 9 days (interquartile range, 3 to 24). Twenty (77%) patients were mechanically ventilated for a median of 13.5 days (interquartile range, 5 to 20). Length of stay and length of mechanical ventilation are likely to be underestimated as 5 patients remain in the hospital (including 2 still in the ICU). Four patients were referred and two were transferred to an extracorporeal membrane oxygenation (ECMO) center. Other adverse ICU outcomes included re-intubation within 48 hours for 1 patient and readmission to the ICU for 2 patients. Six (30%) mechanically ventilated patients were treated for ventilator-associated pneumonia (VAP). One patient developed a catheter-associated urinary tract infection, but no patients developed central-line associated infections.

## Discussion

We report for the first time the results of a case series of critically ill patients with COVID-19 from a region with early public health interventions to slow the spread of SARS-CoV-2 and adequate time and resources to prepare for COVID-19 ICU admissions. We had five weeks to prepare from the time the Mayor declared a state of emergency to our first ICU patient with COVID-19. Although we experienced a higher patient volume than normal, our hospital did not experience any significant shortages of staff, pharmaceuticals, or equipment needed to provide standard ICU management of ARDS. Findings from this case series differ from other published data in two important ways: (1) mortality among critically ill patients with COVID-19, including those mechanically ventilated, has been low, and (2) the lung compliance observed in patients with COVID-19 pneumonia and ARDS has been similar to reported values in patients with ARDS from other causes. Overall, our findings reinforce the importance of public health measures such as physical distancing to enable hospitals to deliver standard care to critically ill patients with respiratory failure from COVID-19 pneumonia.

There are several potential explanations for the lower mortality observed in our cohort compared with previous case series. Patients in this cohort were younger than in previously published case series and were admitted from the community rather than skilled nursing facilities. However, approximately three-quarters had a comorbidity previously associated with worse outcomes in COVID-19 pneumonia and nearly half had two such comorbidities. Our patients were also comparable to other cohorts by standard assessment of severity of illness including length of stay, vasopressor use, need for continuous renal replacement therapy, and APACHE II score.^9^ Despite adherence to lung protective ventilation, a substantial fraction of patients had high driving pressures, a metric associated with mortality in ARDS.^22^ These data, in combination with the use of advanced management for hypoxemic respiratory failure and CRRT in a substantial proportion of patients, argue against the possibility that our low observed mortality reflects a less severely ill population.

Patient care at our institution was focused on proven therapies for ARDS including lung protective ventilation, a fluid conservative strategy, and the use of prone positioning and neuromuscular blockade for moderate to severe disease.^21,23-30^ Adherence to lung protective ventilation for ARDS is known to be generally poor and has not been reported in prior case series of COVID-19 related ARDS.^31^ However, one can infer that in healthcare systems under stress, barriers to implementing these protocols would be even more difficult to overcome. We presume a key driver of good outcomes in this cohort was also our ability to mobilize sufficient critical care trained physician and non-physician staff. Our data suggest that among severely ill patients admitted to the ICU with COVID-19 pneumonia, good outcomes can be achieved so long as healthcare systems are not overwhelmed and are able to deliver evidence-based treatments for ARDS and hypoxemic respiratory failure for prolonged periods. However, clinical trials of experimental treatments to reduce the need for ICU admission, duration of ICU admission and long-term outcomes remain critically important.

Our data suggest that the pulmonary physiology of COVID-19 pneumonia is variable with a range of compliance values similar to ARDS cohorts before the COVID-19 pandemic.^22,32,33^ While the median values of compliance and tidal compliance may be marginally higher than what is reported in studies of ARDS from other causes, we did not observe a preponderance of a high compliance phenotype of COVID-19 ARDS that has been reported elsewhere.^34-36^ We managed our ventilators with standard ARDSNet protocols and did not alter our supportive care strategies for patients with ARDS from COVID-19.

Our study has several limitations. First, this is a small cohort of patients from a single center with censoring of outcome data for 8 patients who remain intubated without adjudication of ultimate disposition. Mortality may be underestimated but would still be at most 40% in ventilated patients, similar to what has been reported for moderate to severe ARDS.^31^ Second, our COVID-19 ICU staffing model and resource mobilization was possible due to monetary support from emergency funds and may not be generalizable in particular to private or non-academically affiliated healthcare systems. Last, our study was not designed to evaluate reasons for the high proportion of Latinx patients in our cohort (77%) compared to the usual patients who receive care in our hospital (25%), an important disparity that requires further study.

Our early experience of the COVID-19 pandemic substantively adds to what is known about outcomes in critically ill patients with COVID-19 pneumonia and does not resemble the experience reported in many other settings. Majority of patients (81%) have been discharged from the ICU, including 65% who have been discharged home. Early efforts to develop coordinated local and regional response plans with contingency planning for personnel, equipment, and pharmaceutical shortages are vital to prepare for later phases of this pandemic with anticipated ebbs and flows in patient volume. While awaiting the results of clinical trials and ultimately an effective vaccine, our findings highlight the ongoing importance of public health measures to minimize peak hospital case load during outbreaks and thereby enable ICUs to deliver standard supportive care for critically ill patients with COVID-19 pneumonia.

## Data Availability

Upon request to the authors, reasonable requests for deidentified data will be accommodated.

## Acknowledgements

We would like to acknowledge the dedication of the administration, clinical staff, trainees, and support staff at ZSFG. We offer gratitude to the elected officials in our region and state as well as the public health officers who acted promptly in response to the COVID-19 pandemic and the members of the public who are following this guidance whenever possible. Dr. Carolyn Hendrickson, MD MPH, MA is supported by a NIH NHLBI Career Development Award, K23 HL133495.

